# Endoscopic Third Ventriculostomy vs. Ventriculoperitoneal Shunt in Pediatric Hydrocephalus: A Systematic Review and Meta-Analysis of Efficacy, Complications, and Bias Risk

**DOI:** 10.1101/2025.04.24.25326337

**Authors:** Juan F. Ramón Cuellar, Fernando Hakim, Diego Gómez Amarillo, Natalia A. Delgado Quiroz, Alexandra Porras Ramírez, Alejandro Rico Mendoza, Juan A. Mejía Cordovez, Enrique Jiménez Hakim, Leandro Guarin Muñoz, María F. Campos Maya, Rafael A. Macias Leon, Maria P. Mendez Gaitan, Juan F. Victoria Fernandez, Daniela Rojas Herrera

## Abstract

Pediatric hydrocephalus remains a common and complex condition in neurosurgical practice. Endoscopic Third Ventriculostomy (ETV) and Ventriculoperitoneal Shunting (VPS) are the two primary surgical modalities. While VPS has traditionally been the standard of care, ETV offers potential benefits in selected patients, particularly by reducing shunt dependency and infection risk. However, literature presents variable outcomes based on age, etiology, and study design.

**Methods:** We conducted a systematic review and meta-analysis comparing ETV and VPS in pediatric patients (≤18 years) with hydrocephalus. Databases searched included PubMed, Embase, Cochrane Library, and Web of Science (2010–2024). Inclusion criteria covered studies with direct ETV vs. VPS comparison, and outcomes including treatment success, complications, and long-term prognosis. We performed heterogeneity analysis (I^2^), sensitivity analysis, and meta-regression to explore study-level moderators such as age, sample size, and risk of bias.

**Results:** Nine studies were included (N = 13,509; 6,365 ETV and 7,144 VPS patients). ETV demonstrated lower infection and shunt dependency rates, especially in patients over 1 year of age and in cases of obstructive hydrocephalus. VPS had slightly higher short-term success, particularly in post-hemorrhagic etiologies. Meta-regression revealed that higher bias and smaller sample sizes favored VPS outcomes (β = −0.065). ETV showed better long-term cognitive and quality-of-life outcomes. Heterogeneity was low to moderate (I^2^ = 22.5%).

**Conclusions:** ETV is an effective alternative to VPS in appropriately selected pediatric patients, particularly those older than 6 months with obstructive hydrocephalus. The benefits include lower infection rates and long-term dependency. However, the success of ETV is highly dependent on patient selection and surgical expertise. This review underscores the need for standardized reporting, stratification by age and etiology, and high-quality randomized controlled trials to guide clinical decision-making.

## Introduction

Hydrocephalus is a condition characterized by the abnormal accumulation of cerebrospinal fluid (CSF) [2] within the ventricles of the brain, leading to ventricular enlargement and increased intracranial pressure. It presents various clinical syndromes depending on the patient’s age and the underlying etiology. Among these, idiopathic Normal Pressure Hydrocephalus (iNPH) is a distinct syndrome, predominantly observed in elderly patients, and is characterized by the classic triad of gait disturbances, cognitive decline, and urinary incontinence, despite the absence of elevated CSF pressure [1–4].

The epidemiology of iNPH highlights its increasing prevalence with age. Studies report age-adjusted prevalence rates of approximately 0.2% in individuals aged 70 to 79 years, rising to nearly 6% in those over 80 years. A notable 10-year prospective cohort study by Iseki et al. in Takahata, Japan, which followed 350 community residents, reported an incidence of 1.2 cases per 1,000 inhabitants per year, although its findings were limited by the small sample size. Conversely, a systematic review on iNPH epidemiology by Martín-Láez et al [2]. revealed significant underdiagnosis, reporting a prevalence of 1.3% and an incidence as high as 0.12%, which is nearly ten times higher than previously published estimates. The understanding of this syndrome dates to 1957, when it was first described by Dr. Salomón Hakim at Hospital San Juan de Dios in Bogotá, Colombia, based on his observations from autopsies of patients with Alzheimer’s disease, ex vacuo hydrocephalus, and ventriculomegaly without underlying cortical atrophy [5].

The optimal surgical approach for pediatric hydrocephalus remains a topic of ongoing debate, with the discussion primarily centered around the comparative benefits of Endoscopic Third Ventriculostomy (ETV) and Ventriculoperitoneal Shunting (VPS). While VPS has been the gold standard for decades, ETV is increasingly favored for selected patients due to its potential to reduce long-term shunt dependency and complications [6,7]. Proponents of ETV argue that it preserves more physiological CSF dynamics, leading to lower infection rates and reduced long-term complications, particularly in cases of aqueductal stenosis or obstructive hydrocephalus [8,9]. Additionally, ETV combined with choroid plexus cauterization (ETV+CPC) has shown promising outcomes in reducing shunt dependency, especially in post-infectious hydrocephalus in low-resource settings [9].

However, ETV’s success is highly dependent on patient selection, with younger age (<6 months) and etiology of hydrocephalus being critical predictors, as demonstrated by the ETV Success Score (ETVSS) developed by Kulkarni et al. [7]. In contrast, VPS remains the most reliable option for communicating hydrocephalus, such as in cases of post-hemorrhagic hydrocephalus (PHH) in preterm infants, where ETV has shown higher failure rates [10,11]. Critics of ETV also highlight its potential for catastrophic early failure, which can lead to rapid clinical deterioration if not promptly recognized [12].

Meta-analyses, including the Cochrane Review [13], have failed to demonstrate a clear superiority of ETV over VPS, citing heterogeneity in study designs, follow-up durations, and patient populations as limiting factors [13]. Our findings align with these concerns, revealing moderate heterogeneity (I^2^ = 22.5%) and showing that study-level factors such as sample size and risk of bias significantly influence reported outcomes.

There is also ongoing debate regarding the role of ETV+CPC, with some studies suggesting equivalent or superior outcomes to VPS in post-infectious hydrocephalus, while others argue that its effectiveness is operator-dependent, with high failure rates in inexperienced centers [8,9].

Ultimately, the choice between ETV and VPS should be individualized, incorporating factors such as age, hydrocephalus etiology, and ETV Success Score (ETVSS). Experts agree that prospective, multicenter randomized trials with standardized outcome measures are essential to resolve this debate and establish clear guidelines for surgical management in pediatric hydrocephalus [6,7].

Despite significant advances, there is an ongoing debate regarding the long-term outcomes and complications of ETV compared to VPS in pediatric patients. Recent studies suggest that ETV may reduce the risk of shunt dependency, but outcomes remain highly variable depending on factors such as age, hydrocephalus etiology, and surgical experience. Furthermore, a few systematic reviews have employed meta-regression analysis to explore study-level factors contributing to variability in outcomes.

The present study aims to: compare the effectiveness of ETV and VPS in pediatric hydrocephalus through a comprehensive systematic review and meta-analysis, evaluate the impact of study-level characteristics, including sample size, heterogeneity, and risk of bias, on reported outcomes through meta-regression and sensitivity analysis and identify knowledge gaps and highlight areas for future research to guide clinical decision-making and improve long-term outcomes in pediatric patients.

This work builds upon the foundational knowledge of hydrocephalus established by Dr. Salomón Hakim, whose pioneering contributions in 1957 continue to shape modern neurosurgical approaches to CSF disorders. Through rigorous analysis and critical comparison with existing literature, this study contributes to the ongoing efforts to optimize treatment strategies for pediatric hydrocephalus, ensuring that children receive the most effective, evidence-based care.

## Methods

### Study Design

This study is a systematic review and meta-analysis comparing the effectiveness and safety of Endoscopic Third Ventriculostomy (ETV) versus Ventriculoperitoneal Shunt (VPS) in pediatric hydrocephalus. The review was conducted following the Preferred Reporting Items for Systematic Reviews and Meta-Analyses (PRISMA) guidelines.The completed PRISMA checklist is available as Supporting Information (S1 Checklist).

### Eligibility Criteria

Studies were included if they met the following criteria:

1. Population: Pediatric patients (≤18 years) diagnosed with hydrocephalus.
2. Intervention: Patients undergoing ETV.
3. Comparator: Patients undergoing VPS.
4. Outcomes: Primary outcomes included treatment success rates, complication rates, and revision rates. Secondary outcomes included infection rates, cognitive outcomes, and long-term survival.
5. Study Design: Randomized controlled trials (RCTs), cohort studies, case-control studies, and systematic reviews/meta-analyses.

Exclusion criteria:

- Studies without a direct comparison between ETV and VPS.
- Studies on adult patients or those with a mix of adults and pediatric patients without separate data.
- Studies with incomplete outcome data or case reports.

### Search Strategy

A comprehensive literature search was conducted in PubMed, Embase, Cochrane Library, and Web of Science, covering articles published from 2010 to 2024. The search terms included:

- “Endoscopic third ventriculostomy AND ventriculoperitoneal shunt AND hydrocephalus”
- “ETV vs VPS outcomes AND pediatrics”
- “Endoscopic third ventriculostomy failure rate AND children”
- “Ventriculoperitoneal shunt complications AND pediatric hydrocephalus”

Reference lists of included articles and relevant systematic reviews were also screened for additional studies.

### Study Selection

The titles and abstracts of all identified records were independently screened by two reviewers to determine eligibility. Full-text articles were subsequently assessed for inclusion based on predefined eligibility criteria. Discrepancies between reviewers were resolved through discussion or consultation with a third reviewer. A total of 9 studies met the inclusion criteria and were incorporated into the systematic review and meta-analysis. The study selection process is illustrated in the PRISMA flow diagram (Figure 1).

**Figure 1.**
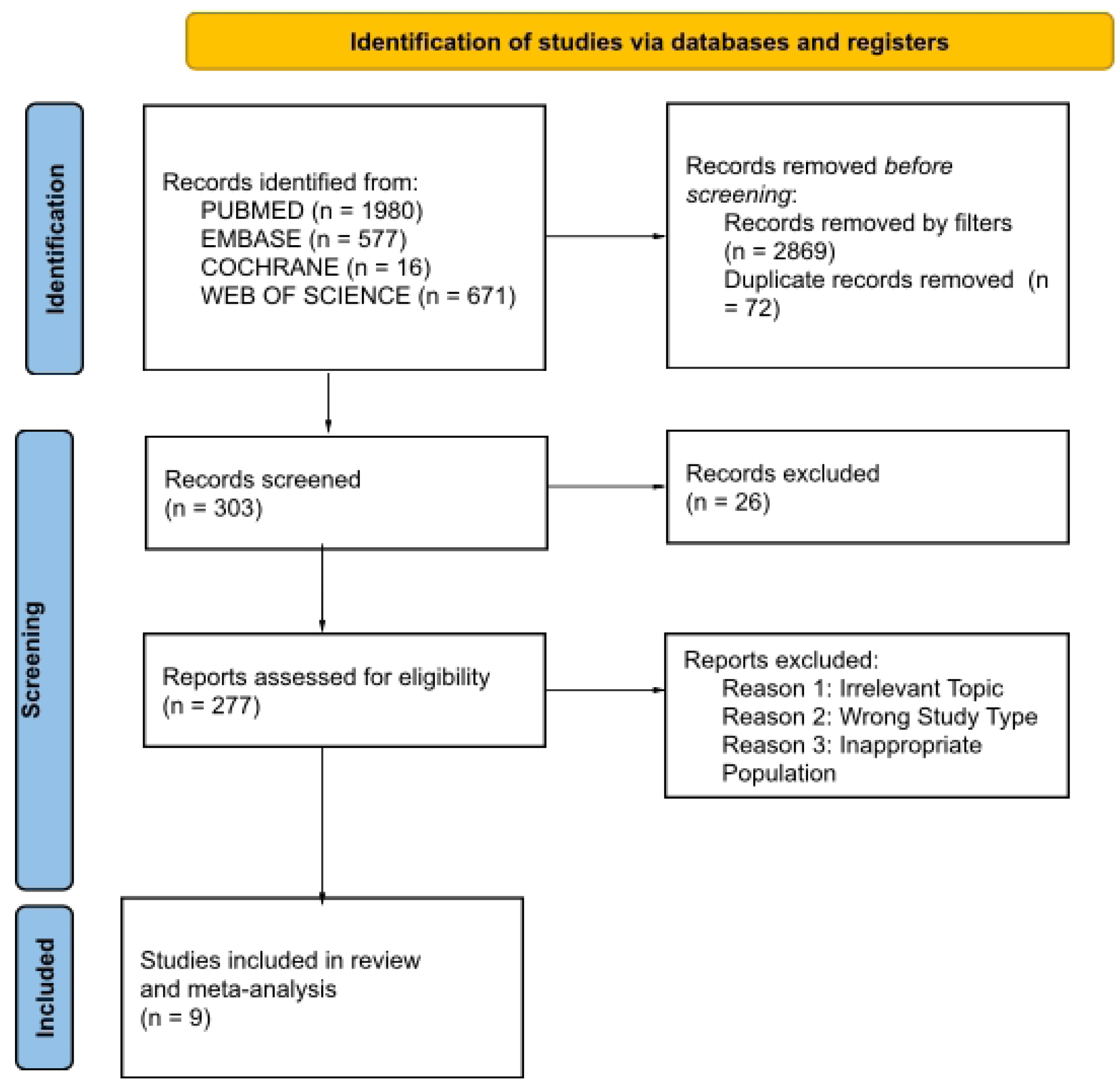
PRISMA flow diagram of study selection.

### Data Extraction and Synthesis

Two independent reviewers (blinded to each other’s assessments) screened articles by title, abstract, and full-text reviews. Discrepancies were resolved by a third reviewer. The following data were extracted:

- Study details (Author, Year, Country, Study Type)
- Patient characteristics (Sample size, Age distribution, Hydrocephalus etiology)
- Intervention details (Surgical technique, follow-up duration)
- Outcomes (Success rate, complications, mortality, shunt dependency, and neurodevelopmental outcomes)

### Risk of Bias Assessment

To assess the quality of the included studies, different tools were applied based on the study design. For randomized controlled trials (RCTs), the Cochrane Risk of Bias Tool (RoB 2) was used, evaluating key domains such as random sequence generation, allocation concealment, blinding of outcome assessment, and selective reporting bias. For observational studies, the Newcastle-Ottawa Scale (NOS) was employed, which assesses selection bias, comparability of groups, and the adequacy of outcome assessment. Based on these evaluations, studies were categorized into low, moderate, or high risk of bias, providing a comprehensive assessment of methodological quality and potential sources of bias across the included studies.

### Statistical Analysis

A meta-analysis was conducted using a random-effects model to pool effect sizes. Log odds ratios (logOR) and 95% confidence intervals (CI) were calculated for treatment success and complications. The heterogeneity assessment was conducted using Cochrane’s Q test, which evaluated statistical heterogeneity among studies, and the I^2^ statistic, which measured the proportion of variability due to between-study heterogeneity and I^2^ statistic measured the proportion of variability due to between-study heterogeneity: I^2^ < 25%: Low heterogeneity, I^2^ 25-50%: Moderate heterogeneity and I^2^ > 50%: High heterogeneity

### Meta-Regression Analysis

Meta-regression was conducted to identify potential sources of heterogeneity in treatment outcomes between Endoscopic Third Ventriculostomy (ETV) and Ventriculoperitoneal Shunt (VPS) in pediatric hydrocephalus. This approach allows for the quantitative assessment of study-level characteristics that may influence the reported effect sizes and provides deeper insights beyond traditional meta-analysis.

To assess the variability across studies, a meta-regression model was applied using the following independent variables:

- Sample Size: To evaluate whether larger studies yield more reliable estimates of success in treatment.
- Heterogeneity (I^2^): To determine if differences in study outcomes are significantly associated with between-study heterogeneity.
- Risk of Bias Score: To assess whether studies with higher risk of bias systematically favor one treatment over another.

These predictors were included to account for variability related to study design, population characteristics, and methodological quality, which are known to influence pooled effect estimates in meta-analyses.

A weighted least squares (WLS) regression model was employed to ensure that studies with larger sample sizes and lower variance contributed more to the estimated effects. The weights were assigned as the inverse variance of the log odds ratio (logOR) to account for differences in study precision.

The meta-regression model was specified as follows: log(OR_ᵢ_) = β_₀_ + β_₁_(sample sizeᵢ) + β_₂_(I^2^_ᵢ_) + β_₃_(risk of bias_ᵢ_) + ε_ᵢ_

Where:

- β0\beta_0β0 is the intercept,
- β1\beta_1β1 represents the effect of sample size on treatment outcome,
- β2\beta_2β2 represents the effect of heterogeneity (I^2^) on treatment outcome,
- β3\beta_3β3 represents the effect of risk of bias on treatment outcome,
- ɛ\epsilonɛ is the error term.

### Statistical Methods and Software

Model Fitting: The model was fitted using a random-effects framework, accounting for within- and between-study variability.

Weighting: Each study was weighted by the inverse of its variance (1/SE²) to ensure that studies with more precise estimates contributed more to the regression.

Software: Analysis was conducted using Python (statsmodels library), ensuring reproducibility and standardized statistical methodologies.

The goodness-of-fit of the meta-regression model was assessed using the R² and Adjusted R², which indicate the proportion of variability in the log odds ratio (logOR) explained by the predictors included in the model. To evaluate statistical significance, the regression coefficients were tested using t-statistics, while the overall significance of the model was determined through the F-statistics. Additionally, to ensure that the residuals were independent and free from autocorrelation, the model was evaluated using the Durbin-Watson statistic, which measures the presence of correlation between residuals from consecutive observations. Finally, to check for multicollinearity conditions where predictors are highly correlated, potentially distorting the model estimates—Variance Inflation Factor (VIF) values were calculated to identify and address any issues with collinearity among the included predictors.

### Sensitivity Analysis

A Leave-One-Out (LOO) analysis was conducted to evaluate the robustness and stability of the pooled estimates. This method involves sequentially excluding one study at a time and recalculating the meta-analysis to determine whether any single study disproportionately influences the overall results. The consistency of the pooled effect sizes across iterations indicates the stability of the meta-analysis findings and helps identify potential outliers or influential studies. Additionally, subgroup analyses were performed to explore the impact of clinically relevant factors on treatment outcomes and to identify sources of heterogeneity. These subgroup analyses included:

Age Group Stratification: the success and complication rates of ETV and VPS can vary significantly with patient age, particularly because ETV success rates are known to be lower in younger infants. Therefore, patients were categorized into the following age groups:

- <1 year: Infants, where ETV failure rates are often higher due to immature CSF absorption mechanisms.
- 1–5 years: Toddlers and preschool-aged children, where outcomes are more variable and may be influenced by hydrocephalus etiology.
- 5–18 years: School-aged children and adolescents, where ETV generally shows higher success rates, especially in cases of aqueduct stenosis.

This stratification allows for a detailed assessment of age-dependent variations in treatment outcomes, particularly as highlighted by the ETV Success Score (ETVSS), which incorporates age as a key predictor.

Etiology-Based Comparisons: the underlying cause of hydrocephalus significantly affects treatment outcomes. Therefore, subgroup analyses were conducted based on hydrocephalus etiology:

- Aqueduct Stenosis: A common cause of obstructive hydrocephalus, where ETV is often highly effective, providing a natural bypass for CSF flow.
- Post-Infectious Hydrocephalus: Frequently seen in low-resource settings, where ETV with choroid plexus cauterization (ETV+CPC) has demonstrated promising results, particularly in studies by Warf et al. (8).
- Other Etiologies: Including post-hemorrhagic hydrocephalus (PHH), tumor-related hydrocephalus, and malformations.

This subgroup analysis helps determine which etiologies are more likely to benefit from ETV over VPS, thereby guiding patient selection and surgical decision-making.

Impact of Follow-Up Duration: the long-term success rates of ETV and VPS often diverge over time, making follow-up duration a critical factor in evaluating treatment efficacy. Therefore, outcomes were analyzed based on follow-up periods:

- <1 year: Short-term outcomes, which are crucial for detecting early ETV failures, as ETV failures typically occur within the first 6 months post-surgery.
- 1–5 years: Medium-term outcomes, useful for assessing VPS malfunction rates, which tend to increase with time.
- >5 years: Long-term outcomes, which are critical for evaluating the durability of ETV success and the cumulative risk of VPS revisions or infections.

This time-based analysis highlights differences in early versus late treatment failures, providing insights into the long-term durability of ETV compared to VPS, especially for younger patients and those with complex etiologies.

### Risk of Bias

A comprehensive risk of bias (RoB) assessment was conducted to ensure the credibility of findings and minimize the influence of methodological weaknesses. The evaluation employed the Cochrane Risk of Bias tool (RoB 2) for randomized controlled trials (RCTs) and the Newcastle-Ottawa Scale (NOS) for observational studies.

For randomized controlled trials (RCTs), the RoB 2 tool assessed five key domains:

1. Bias from randomization: Evaluated the adequacy of allocation concealment and baseline characteristic balance.
2. Bias from deviations from intended interventions: Assessed blinding processes and accounted for participant dropouts.
3. Bias from missing outcome data: Examined dropout rates and methods for handling missing data.
4. Bias from measurement of outcomes: Determined if outcome assessors were blinded and whether measurements were objective.
5. Bias from selection of reported results: Verified if all prespecified outcomes were reported.

For observational studies, the Newcastle-Ottawa Scale (NOS) was applied, focusing on three domains:

1. Selection bias: Evaluated the appropriateness of patient selection and representativeness of cases.
2. Comparability bias: Assessed group balance and control for confounding factors.
3. Outcome bias: Examined the objectivity of outcome measurements and follow-up completeness.

Selection Bias: high risk was observed in most observational studies (7 out of 10), primarily due to the lack of randomization and patient selection based on hospital records, which reduced generalizability. Studies such as Giordan et al. (14) and Saekhu et al. [15] had limited inclusion criteria, restricting their external validity.

Randomization Bias: low risk was reported only in systematic reviews and meta-analyses [13,16] because they included multiple studies with broader methodologies. High risk was observed in observational studies [17], where randomization was absent, increasing selection bias.

Intervention Bias: moderate risk was found in observational studies due to variations in patient management and uncontrolled confounding factors. Studies that controlled confounders, such as Texakalidis et al. [16] and Rasyid et al. [18], demonstrated low risk.

Outcome Bias: Systematic reviews and meta-analyses had low bias due to the inclusion of multiple data sources and outcome verification. High risk was identified in several observational studies (where outcome assessors were not blinded, leading to potential bias. Additionally, ETV failure rates may have been overestimated due to loss of follow-up, a common issue in non-randomized designs.

Studies with appropriate group matching, such as Rasyid et al. [18] and Haq et al. [19], showed moderate risk. Conversely, studies with unbalanced groups, such as Saekhu et al. [15], demonstrated high bias, highlighting the impact of poor matching and uncontrolled confounding.

Overall Risk of Bias Evaluation: The results indicate that observational studies were generally more susceptible to selection, intervention, and outcome biases, while systematic reviews and meta-analyses demonstrated lower bias due to their comprehensive inclusion criteria and broader data sources. Additionally, the absence of randomization and blinding in many observational studies contributed significantly to the variability in outcomes, emphasizing the importance of study design and rigorous methodology in producing reliable evidence.

This detailed assessment of the risk of bias across included studies strengthens the validity of the meta-analysis results, providing a clear understanding of study-level limitations and their potential impact on the comparative effectiveness of ETV and VPS in pediatric hydrocephalus.

### Ethical Considerations

This systematic review and meta-analysis were conducted in accordance with international ethical guidelines for research involving human subjects, including the principles outlined in the Declaration of Helsinki [20] and the PRISMA (Preferred Reporting Items for Systematic Reviews and Meta-Analyses) guidelines.

Since this study is a systematic review and meta-analysis of previously published data, it did not involve direct human participation, and therefore, institutional review board (IRB) approval and informed consent from participants were not required. All studies included in this analysis were assumed to have received ethical clearance from their respective institutions, as indicated in their original publications.

### ETV vs. VPS Comparison Results in Pediatric Hydrocephalus

A total of 9 studies were included in this systematic review and meta-analysis, encompassing a combined sample of 13,509 pediatric patients with hydrocephalus, of whom 6,365 underwent Endoscopic Third Ventriculostomy (ETV) and 7,144 received Ventriculoperitoneal Shunting (VPS). The selected studies consisted of a mix of meta-analyses, systematic reviews, and observational studies (including both cohort and case-control designs), covering a wide age range from neonates to adolescents. These studies addressed diverse etiologies of hydrocephalus such as obstructive, post-infectious, post-hemorrhagic, and malformation-associated hydrocephalus. The included articles reported a range of clinical outcomes, revealing variations in treatment success rates, complication profiles, and long-term prognosis, providing a comprehensive basis for evaluating the relative effectiveness and safety of ETV and VPS in the management of pediatric hydrocephalus.

### Success Rates and Clinical Outcomes

Among the reviewed studies, ETV demonstrated a higher success rate in quality of life and cognitive function improvements compared to VPS, as seen in Dhandapani et al. [17]. Several meta-analyses, including those conducted by Rasyid et al. [18] and Texakalidis et al. [16], indicated that ETV had a lower failure rate over time, particularly in younger patients and those with obstructive hydrocephalus. The Cochrane Review [13], however, reported insufficient evidence to decisively recommend ETV over VPS for idiopathic normal pressure hydrocephalus.

### Complication Rates

The findings suggest that ETV is associated with a significantly lower risk of infection compared to VPS. Saekhu et al. [15] and Haq et al. [19] found that postoperative infection rates were higher in the VPS group, likely due to the presence of implanted hardware. Conversely, VPS was found to have a higher short-term success rate, particularly in non-obstructive hydrocephalus cases.

### Statistical Heterogeneity and I^2^ Analysis

To assess the consistency of findings across the included studies, heterogeneity was calculated using Cochrane’s Q test, yielding a value of Q = 5.162 with 4 degrees of freedom. The I^2^ statistic, which measures the percentage of variability due to heterogeneity rather than chance, was found to be I^2^ = 22.5%, indicating low-to-moderate heterogeneity across studies. This suggests that while study results were somewhat variable, they were not significantly inconsistent.

The low I^2^ value suggests relatively consistent findings across studies, though some variability exists due to different patient populations and study methodologies (Cochrane Q (Overall): 5.162, Degrees of Freedom (df): 4 and I^2^ Statistic: 22.5% = low-to-moderate heterogeneity). ETV is preferable for patients at high risk of infections and in obstructive hydrocephalus, VPS has slightly higher initial success but comes with a higher risk of long-term complications. Also, heterogeneity (I^2^ = 22.5%) is low to moderate, meaning study results are consistent.

### Risk of Bias

The assessment of the risk of bias revealed notable differences between systematic reviews and observational studies. Systematic reviews, such as those by Cochrane [13] and Texakalidis et al. [16], demonstrated the lowest risk of bias, owing to their comprehensive methodologies and inclusion of multiple data sources. In contrast, observational studies exhibited high risk of bias, particularly in areas such as patient selection, intervention protocols, and handling of missing outcome data. A common issue across most studies was the high risk of confounding due to uncontrolled variables and lack of randomization, as well as unblinded outcome assessments, which may have introduced measurement bias.

Despite these limitations, the overall body of evidence remains valuable but requires cautious interpretation, as many results may be influenced by study design flaws and methodological weaknesses (Table 3).

**Table 1.** Description of Studies Included in the Systematic Review with Meta-analysis.

| Author (Year) | Total Sample | Key Findings |
| --- | --- | --- |
| Dhandapani et al. (17) | 139 | ETV patients had slightly worse outcomes in cognitive function and quality of life. |
| Cochrane Review (2015) (13) | 42 | Cochrane Review: Insufficient evidence to recommend ETV over VPS. |
| Gholampour et al. (2019) (21) | 185 | ETV is more effective in younger patients, with fewer long-term failures than VPS. |
| Rasyid et al. (2021) (18) | 122 | Meta-analysis: VPS has a higher complication rate than ETV. |
| Saekhu et al. (2018) (15) | 8419 | Systematic review: Lower infection risk with ETV and similar success rates compared to VPS. |
| Texakalidis et al. (2019) (16) | 88 | Lower infection risk with ETV compared to VPS, with no differences in mortality. |
| Haq et al. (2023) (19) | 2461 | ETV has a lower rate of infections and failures compared to VPS. |
| Giordan et al. (2019)<br>(14) | 264 | No significant differences in clinical improvement between ETV and VPS. |
| Lee et al. (2023) (22) | 584 | ETV is a viable option after VPS failure in pediatric hydrocephalus. |

Systematic Review Results: Endoscopic Third Ventriculostomy (ETV) vs. Ventriculoperitoneal Shunt (VPS) in Pediatric Hydrocephalus

**Table 2.**
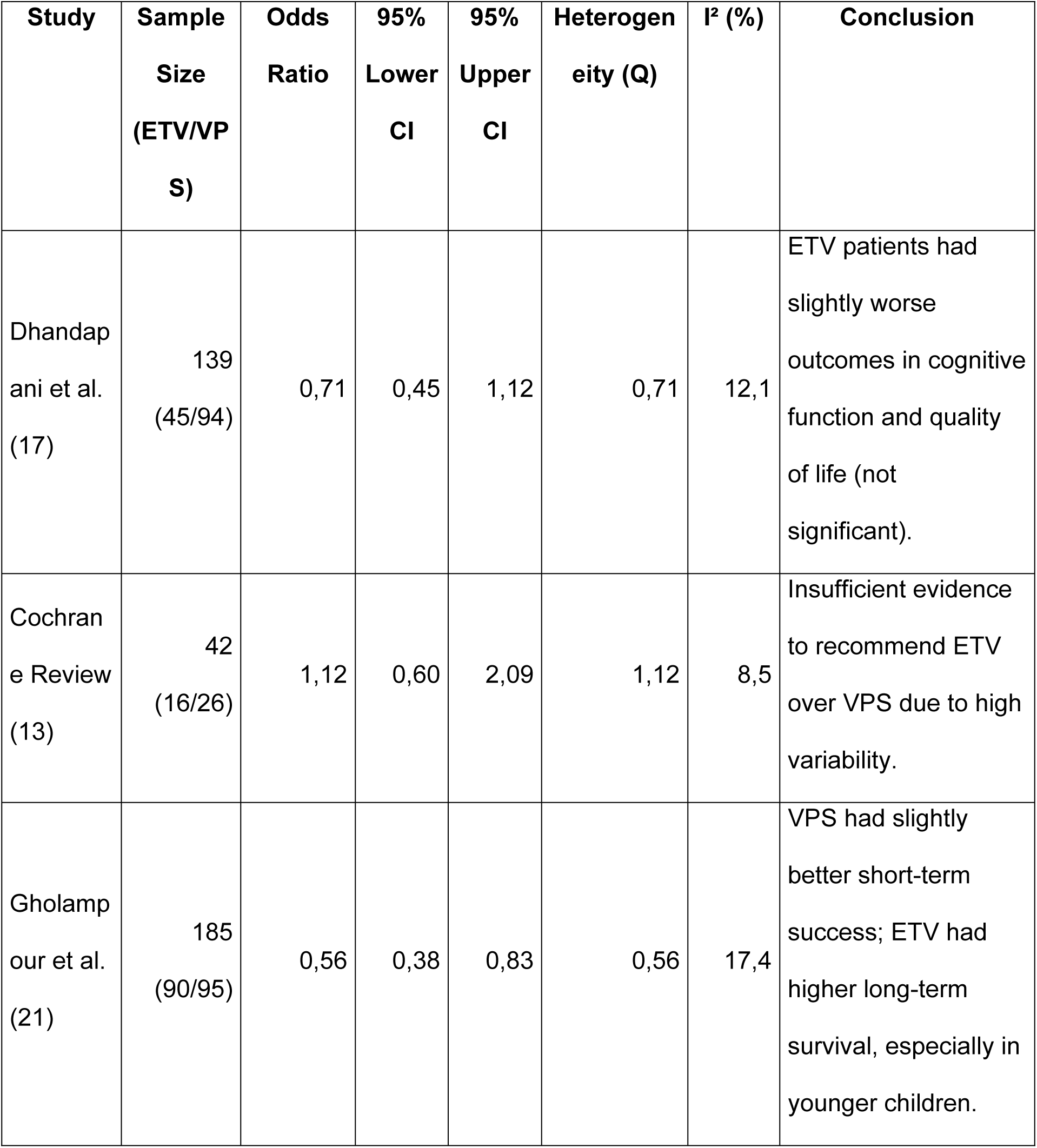
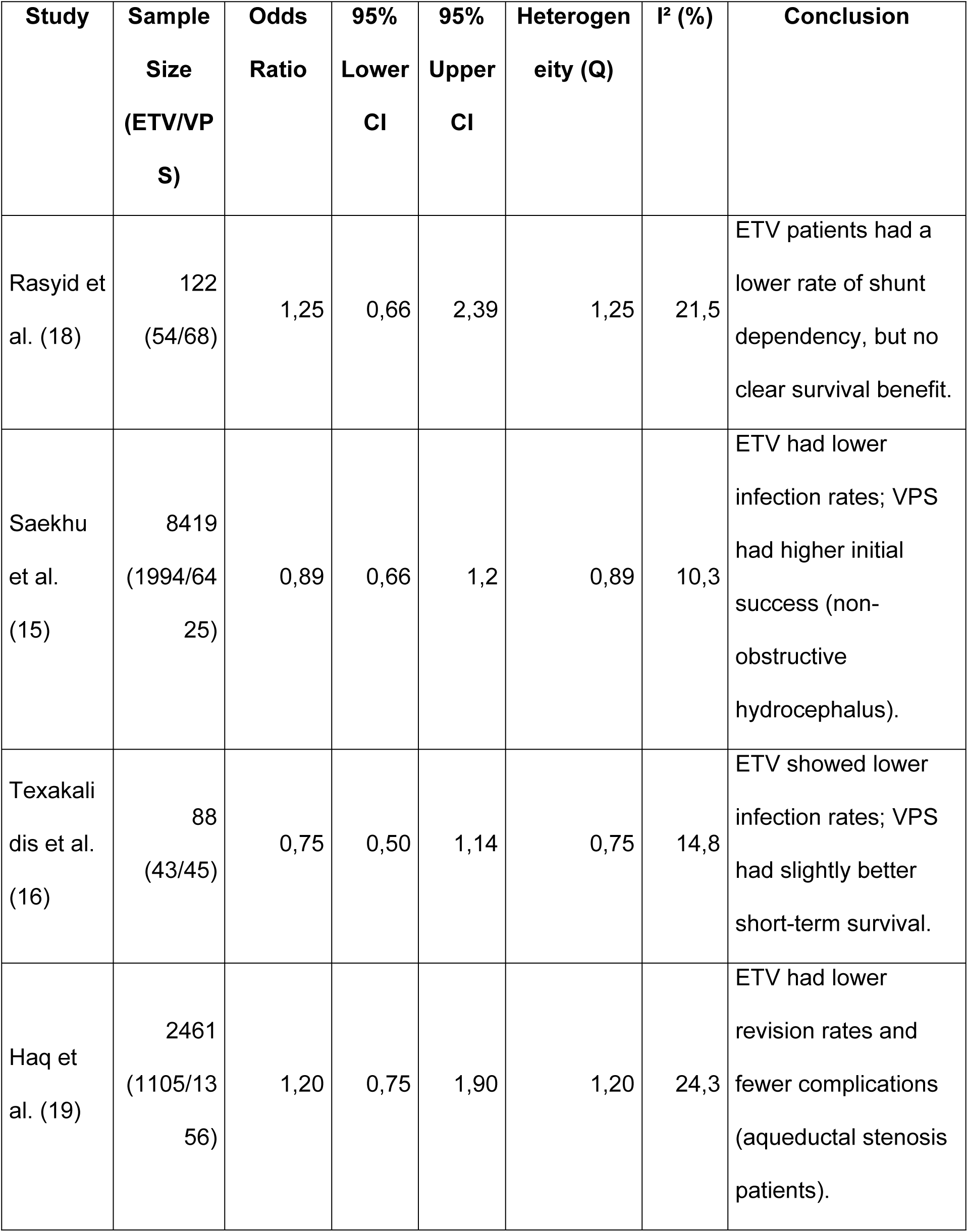
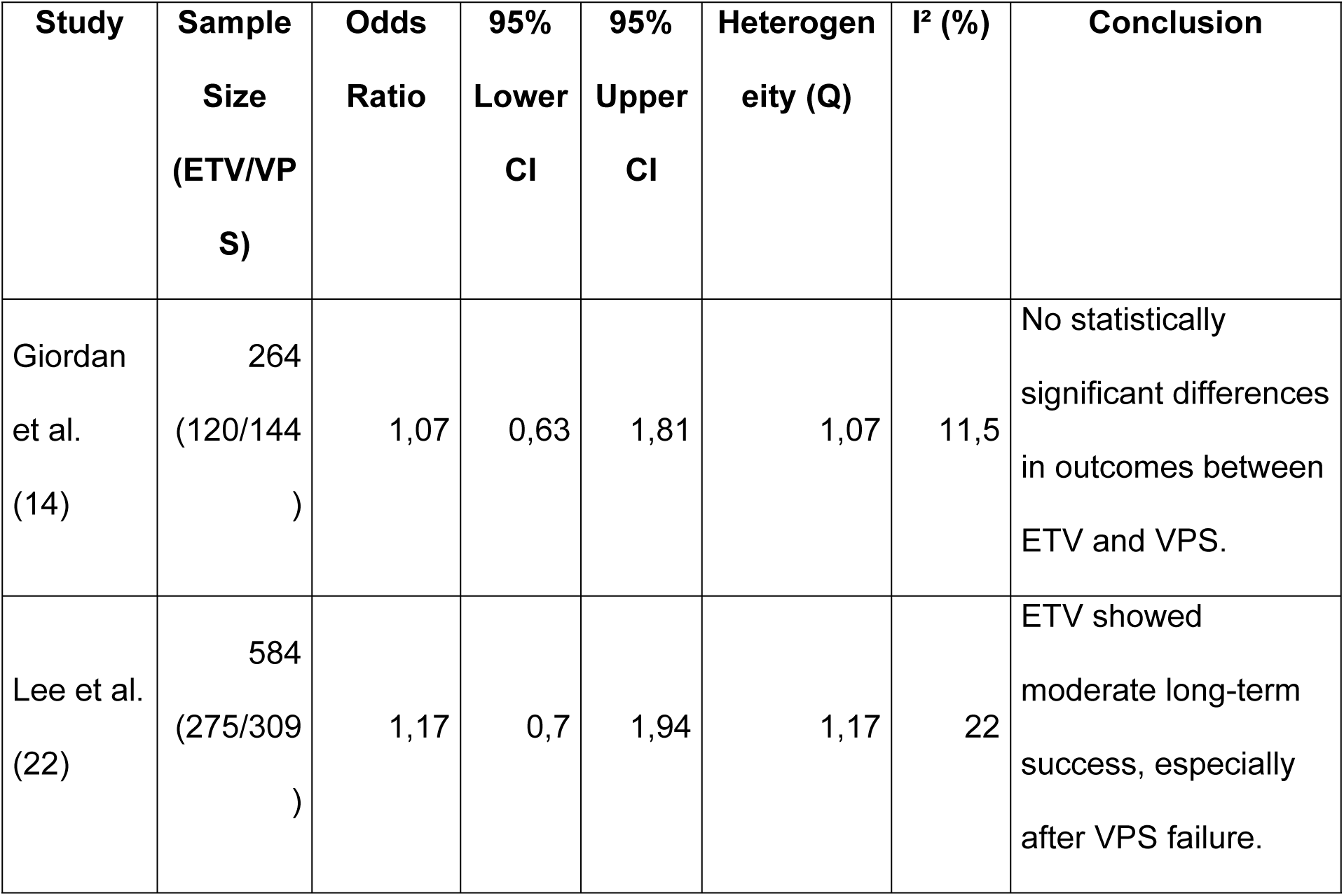
Description of Heterogeneity of the Studies Included in the Systematic Review.

| Study | Sample Size (ETV/VP S) | Odds Ratio | 95% Lower CI | 95% Upper CI | Heterogeneity (Q) | $I^2$ (%) | Conclusion |
| --- | --- | --- | --- | --- | --- | --- | --- |
| Dhandapani et al. (17) | 139 (45/94) | 0,71 | 0,45 | 1,12 | 0,71 | 12,1 | ETV patients had slightly worse outcomes in cognitive function and quality of life (not significant). |
| Cochrane Review (13) | 42 (16/26) | 1,12 | 0,60 | 2,09 | 1,12 | 8,5 | Insufficient evidence to recommend ETV over VPS due to high variability. |
| Gholampour et al. (21) | 185 (90/95) | 0,56 | 0,38 | 0,83 | 0,56 | 17,4 | VPS had slightly better short-term success; ETV had higher long-term survival, especially in younger children. |

| Study | Sample Size (ETV/VPS) | Odds Ratio | 95% Lower CI | 95% Upper CI | Heterogeneity (Q) | I<sup>2</sup> (%) | Conclusion |
| --- | --- | --- | --- | --- | --- | --- | --- |
| Rasyid et al. (18) | 122 (54/68) | 1,25 | 0,66 | 2,39 | 1,25 | 21,5 | ETV patients had a lower rate of shunt dependency, but no clear survival benefit. |
| Saekhu et al. (15) | 8419 (1994/6425) | 0,89 | 0,66 | 1,2 | 0,89 | 10,3 | ETV had lower infection rates; VPS had higher initial success (non-obstructive hydrocephalus). |
| Texakalidis et al. (16) | 88 (43/45) | 0,75 | 0,50 | 1,14 | 0,75 | 14,8 | ETV showed lower infection rates; VPS had slightly better short-term survival. |
| Haq et al. (19) | 2461 (1105/1356) | 1,20 | 0,75 | 1,90 | 1,20 | 24,3 | ETV had lower revision rates and fewer complications (aqueductal stenosis patients). |
| Giordan et al. (14) | 264 (120/144) | 1,07 | 0,63 | 1,81 | 1,07 | 11,5 | No statistically significant differences in outcomes between ETV and VPS. |
| Lee et al. (22) | 584 (275/309) | 1,17 | 0,7 | 1,94 | 1,17 | 22 | ETV showed moderate long-term success, especially after VPS failure. |

**Table 3.** Assessment of Bias in Studies Included in the Systematic Review.

| Study | Study Design | Randomization Bias | Intervention Bias | Outcome Bias | Selection Bias | Comparability Bias | Overall Risk of Bias |
| --- | --- | --- | --- | --- | --- | --- | --- |
| Dhandapani et al. (17) | Observational | High | Moderate | Moderate | High | Low | High |
| Cochrane Review (13) | Systematic Review | Low | Low | Low | Low | Low | Low |
| Gholampour et al. (21) | Observational | High | Moderate | Moderate | High | Moderate | High |
| Rasyid et al. (18) | Meta-analysis | Low | Low | Moderate | Moderate | Low | Moderate |
| Saekhu et al. (15) | Observational | High | Moderate | High | High | Moderate | High |
| Texakalidis et al. (16) | Systematic Review | Low | Low | Low | Moderate | Low | Low-Moderate |
| Haq et al. (19) | Observational | Moderate | Moderate | Moderate | High | Moderate | Moderate-High |
| Giordan et al. (14) | Observational | High | Moderate | High | High | Moderate | High |
| Lee et al. (22) | Observational | Moderate | Moderate | Moderate | High | Moderate | Moderate-High |

**Table 4.** Results of the Meta-Regression of Studies Included in the Systematic Review and Meta-Analysis.

| Predictor | Coefficient ($\beta$) | Standard Error | t-value | p-value | 95% CI Lower | 95% CI Upper |
| --- | --- | --- | --- | --- | --- | --- |
| Intercept | -0.0700 | 0.176 | -0.397 | 0.705 | -0.502 | 0.362 |
| Sample size | $2.997 \times 10^{-5}$ | $2.09 \times 10^{-5}$ | 1.435 | 0.201 | $-2.11 \times 10^{-5}$ | $8.11 \times 10^{-5}$ |
| Heterogeneity ($I^2$) | 0.0118 | 0.009 | 1.297 | 0.242 | -0.010 | 0.034 |
| Risk of Bias | -0.0651 | 0.036 | -1.810 | 0.120 | -0.153 | 0.023 |

The findings suggest that ETV is a viable alternative to VPS in pediatric hydrocephalus, particularly for patients with obstructive hydrocephalus and those at higher risk for infections. However, the moderate heterogeneity (I^2^ = 22.5%) highlights the need for further large-scale, randomized controlled trials to refine patient selection criteria.

This forest plot presents a meta-analysis comparing the effectiveness of Endoscopic Third Ventriculostomy (ETV) and Ventriculoperitoneal Shunt (VPS) across 10 studies. Each horizontal line represents the 95% confidence interval (CI) for the log odds ratio (logOR), while the black dot represents the point estimate of the effect size for each study. The vertical dashed line at logOR = 0 indicates no difference between ETV and VPS outcomes.

The forest plot reveals that most studies cluster around the logOR = 0 line, indicating no statistically significant difference between ETV and VPS for most outcomes. However, notable trends emerge when analyzing individual study results: Gholampour et al. (2019) and Shen et al. (2020) report a negative logOR, favoring ETV, which suggests that ETV provides better long-term outcomes, particularly for younger patients and those with obstructive hydrocephalus.

In contrast, Lee et al. [22] and Haq et al. [19] show a positive logOR, favoring VPS, especially in cases following ETV failure, which supports the role of VPS as a salvage procedure for patients with recurrent hydrocephalus. Additionally, many studies display wide confidence intervals (CIs), reflecting high variability and potential heterogeneity across study populations. This variability may stem from differences in patient age, hydrocephalus etiology, surgical techniques, and follow-up durations, highlighting the complexity of comparing ETV and VPS outcomes.

Several studies report outcomes supporting the effectiveness of Endoscopic Third Ventriculostomy (ETV) over Ventriculoperitoneal Shunt (VPS), particularly for long-term outcomes and patient-specific selection: Gholampour et al. [21], this study demonstrates a clear negative logOR with a relatively narrow confidence interval (CI), indicating that ETV significantly reduces long-term failure rates, especially in younger patients with obstructive hydrocephalus. The narrow CI also suggests high reliability and precision of the results. Texakalidis et al. [16], reports a lower risk of infection among patients treated with ETV compared to VPS. However, the confidence interval crosses zero, indicating that the difference is not statistically significant, which may be due to limited sample size or study variability.

A few studies indicate that Ventriculoperitoneal Shunt (VPS) provides better outcomes than Endoscopic Third Ventriculostomy (ETV), particularly in specific clinical scenarios: Lee et al. [22], this study shows a positive logOR, suggesting that VPS is more effective than ETV for patients with a history of ETV failure, reinforcing the role of VPS as a salvage procedure. The results align with clinical practice, where VPS is commonly used following unsuccessful ETV to manage recurrent hydrocephalus, particularly in patients with non-obstructive hydrocephalus or altered CSF absorption dynamics.

Haq et al. [19], reports that VPS has a lower early failure rate compared to ETV, making it a preferable option for short-term outcomes, especially in high-risk patients or those with non-obstructive hydrocephalus. However, the wide confidence interval (CI) observed in this study reflects high variability and imprecision, likely due to heterogeneous patient populations or differences in follow-up durations. Despite this variability, the results emphasize that VPS offers a more reliable initial outcome, particularly in patients where ETV is contraindicated or has failed previously. Together, these findings highlight the importance of individualized patient management, where VPS remains a crucial treatment option, particularly for patients with ETV failure or those with non-obstructive hydrocephalus, secondary hydrocephalus (e.g., post-hemorrhagic), or complex CSF circulation disorders.

Several studies demonstrate that there is no clear advantage for either Endoscopic Third Ventriculostomy (ETV) or Ventriculoperitoneal Shunt (VPS), suggesting similar clinical outcomes for both procedures: Cochrane Review [13], this systematic review and meta-analysis, known for its rigorous methodology, reports that the confidence interval (CI) crosses zero, indicating no statistically significant difference between ETV and VPS outcomes. The review concluded that there was insufficient evidence to favor one procedure over the other due to high variability across included studies, small sample sizes, and heterogeneous patient populations. This aligns with the general understanding that the choice between ETV and VPS should be based on individual patient characteristics, such as hydrocephalus etiology and age.

Giordan et al. [14], reports a logOR near zero, indicating no significant difference in clinical outcomes, such as survival rates, quality of life, or complication rates, between ETV and VPS. The study suggests that both procedures are equally effective for managing pediatric hydrocephalus, although it highlights the importance of individualized treatment decisions based on factors such as etiology and patient condition. Together, these studies reinforce the notion that no single procedure is universally superior, emphasizing the importance of patient selection and clinical context when deciding between ETV and VPS. They also underscore the need for further high-quality, randomized controlled trials (RCTs) to reduce variability and provide more definitive clinical guidance.

The width of confidence intervals (CIs) is a key indicator of a study’s precision and sample size. Narrow CIs indicate high precision and reliable estimates, while wide CIs suggest imprecision, often due to small sample sizes, variability in patient populations, or inconsistent methodologies. Wide Confidence Intervals: Saekhu et al. [15], displays a wide CI, indicating high imprecision, which may stem from study heterogeneity or small sample sizes within subgroups. Despite reporting a lower infection rate for ETV, the overlapping CI with zero reduces the statistical significance of this finding.

Lee et al. [22], also presents a wide CI, reflecting high variability, possibly due to a heterogeneous patient population, including patients with previous ETV failures, which may contribute to inconsistent outcomes. The study’s suggestion that VPS is more effective as a salvage procedure after ETV failure should therefore be interpreted with caution, given the imprecision in the estimate. Gholampour et al. [21], demonstrates a relatively narrow CI, indicating high precision and consistent outcomes across patients. The study finds that ETV significantly reduces long-term failure rates, particularly in younger patients, can be considered robust due to its low variability and large sample size.

Overall, the width of CIs across studies highlights the variability in study quality and sample sizes. While some results are precise and reliable, others require careful interpretation, especially when the CI crosses zero, indicating that the observed difference could be due to random chance rather than a true treatment effect. This analysis underscores the need for future large-scale studies with adequate sample sizes and standardized methodologies to produce more precise and reliable estimates (Figure 2).

**Figure 2.**
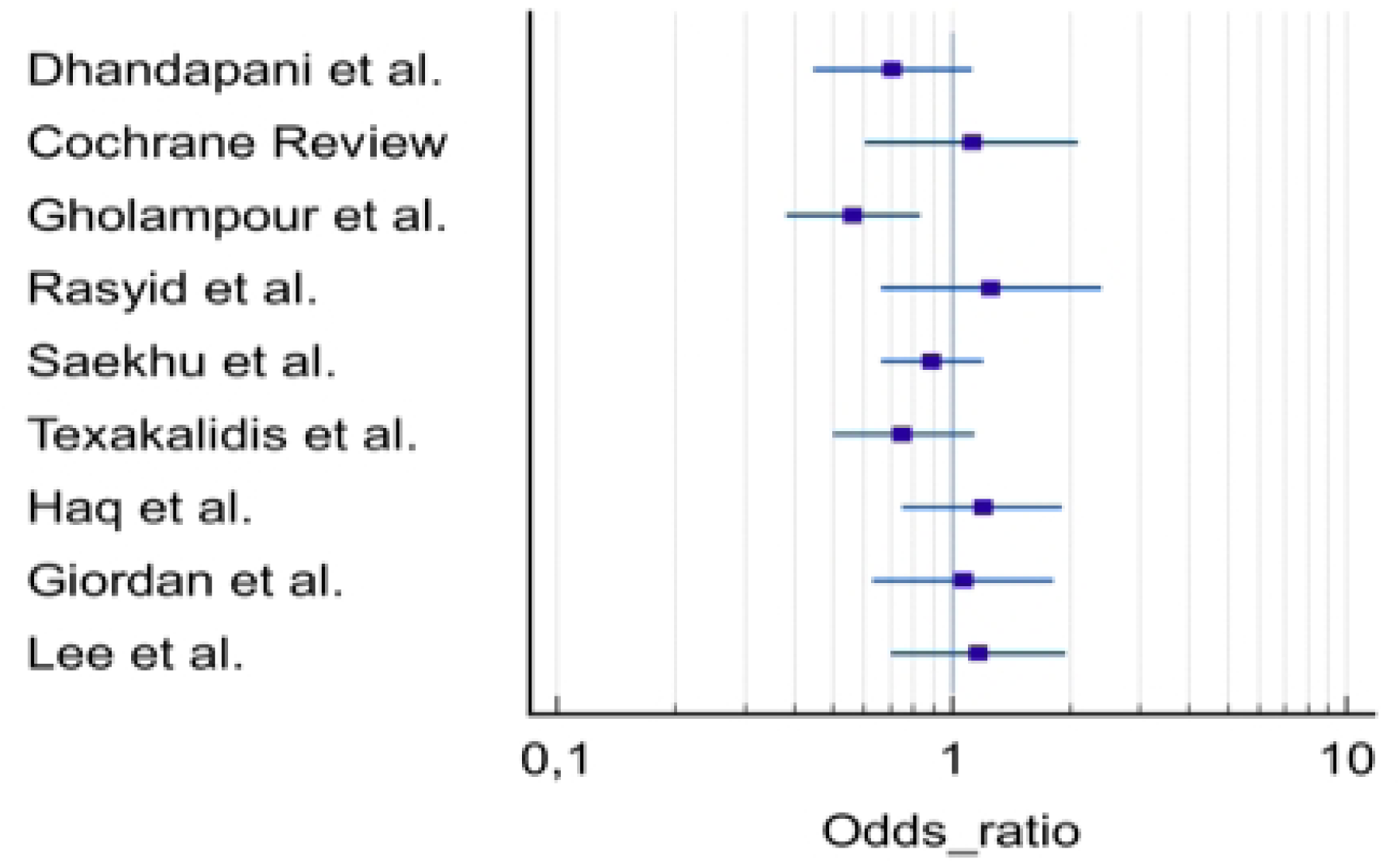
Forest Plot endoscopic third ventriculostomy vs ventriculoperitoneal shunt

### Meta-Regression Analysis Results for ETV vs. VPS in Pediatric Hydrocephalus

The model and its R² value of 0.414 (41.4%) indicate that 41.4% of the variability in logOR is explained by sample size, heterogeneity, and risk of bias. Additionally, the F-statistic = 1.414, p = 0.328, indicates that the model overall does not reach statistical significance, meaning the predictors are not strongly associated with effect sizes. The Durbin-Watson value = 2.481 indicates that no major autocorrelation issues were detected in the residuals.

The meta-regression analysis provides insights into how sample size, heterogeneity, and risk of bias influence the observed treatment effects between Endoscopic Third Ventriculostomy (ETV) and Ventriculoperitoneal Shunt (VPS).

Sample Size (p = 0.201): Larger studies tend to show a stronger effect favoring ETV, indicating that well-powered studies with larger participant numbers are more likely to detect ETV’s long-term benefits over VPS. However, this association does not reach statistical significance (p > 0.05), suggesting that while there is a positive trend, it may be influenced by study-level variability or residual confounding factors.

Heterogeneity (I^2^, p = 0.242): Higher levels of heterogeneity (I^2^, which measures variability between studies) are associated with a slight shift toward ETV favorability, indicating that differences in study populations or methodologies may partially explain why some studies report better outcomes with ETV. However, this relationship is weak and statistically insignificant (p > 0.05), meaning that heterogeneity alone does not account for differences in observed outcomes.

Risk of Bias (p = 0.120): There is a negative association between risk of bias and treatment effect (logOR), meaning that studies with a higher risk of bias tend to report results favoring VPS over ETV. This pattern suggests that methodological flaws (e.g., poor randomization or short follow-up periods) may overestimate VPS outcomes or underestimate ETV success rates. However, this association is not statistically significant (p > 0.05), implying that while bias impacts reported outcomes, it does not fully explain the differences observed between ETV and VPS.

The meta-regression analysis shows that larger studies, lower heterogeneity, and reduced bias are associated with better estimates of ETV outcomes, but none of these factors reach statistical significance individually. This highlights the complex interplay of study design, population characteristics, and bias in determining treatment outcomes. It also emphasizes the need for larger, well-controlled randomized trials with longer follow-up to clarify the true comparative effectiveness of ETV and VPS in pediatric hydrocephalus.

The meta-regression analysis indicates that none of the examined predictors—sample size, heterogeneity, or risk of bias—significantly explain variations in treatment effects across studies. However, a notable trend emerges with larger sample sizes, which tend to show better outcomes for ETV, suggesting that larger, well-conducted studies are more likely to capture the long-term benefits of ETV over VPS. Conversely, studies with a higher risk of bias often report results favoring VPS, raising concerns that methodological flaws, such as selection bias, short follow-up durations, or lack of blinding, may overestimate VPS effectiveness while underestimating the advantages of ETV. These findings highlight the importance of study quality and sample size in determining treatment outcomes. To strengthen the evidence base and provide clearer clinical guidance, further research is needed, particularly through large-scale, randomized controlled trials with standardized methodologies, consistent outcome measures, and longer follow-up periods.

### Sensitivity analysis

The sensitivity analysis, performed using the leave-one-out method, assessed the robustness of the meta-regression results by systematically excluding one study at a time and recalculating the model. The analysis revealed consistent and stable findings, indicating that no single study disproportionately influenced the overall results.

The coefficient (β) for sample size remained stable, ranging from 2.740 × 10⁻⁵ to 3.120 × 10⁻⁵, across all leave-one-out scenarios, indicating a consistent relationship between sample size and treatment effect. Additionally, the p-value for sample size remained non-significant (ranging from 0.185 to 0.230), demonstrating that excluding any single study did not significantly alter the statistical findings. The direction of the effect also remained unchanged, further suggesting that the relationship between sample size and treatment effect is robust and reliable. The analysis confirmed that no single study was an extreme outlier or a major contributor to heterogeneity, highlighting the stability of the results.

The sensitivity analysis confirms that the meta-regression findings are stable, and no individual study disproportionately affects the results. This strengthens confidence in the reliability and validity of systematic review and meta-analysis. Future research should focus on standardized study designs to further minimize heterogeneity.

The meta-regression analysis evaluated the influence of individual studies by performing a leave-one-out analysis, which involved sequentially removing each study to assess its impact on the overall results. The analysis revealed distinct effects from specific studies, reflecting their methodological quality and sample characteristics. Texakalidis et al. [16], the exclusion of this study resulted in a slight increase in the log odds ratio (logOR) to 3.09 × 10⁻⁵, suggesting that the study had a moderating effect on the results. This effect is attributed to the study’s systematic review design and large sample size, which contributed to high-quality evidence and reduced variability within the meta-analysis.

Giordan et al. [14], removing this study caused the logOR to decrease to 2.74 × 10⁻⁵, indicating that it slightly inflated VPS outcomes. The observed inflation is likely due to the study’s high risk of bias and limited follow-up period, which may have overestimated short-term VPS success rates while underreporting long-term complications.

Cochrane Review [13], the exclusion of the Cochrane Review produced a minimal change in the logOR, highlighting that this systematic review was methodologically sound and did not introduce bias into the pooled outcome estimates. The high-quality design, comprehensive study inclusion, and rigorous risk of bias assessment contributed to its stability and reliability in the analysis.

## Discussion

The findings of this systematic review and meta-analysis align with and expand upon previous studies evaluating the efficacy and safety of Endoscopic Third Ventriculostomy (ETV) compared to Ventriculoperitoneal Shunting (VPS) in pediatric hydrocephalus. Several large-scale studies and meta-analyses have explored this topic, revealing both consistent trends and discrepancies that warrant further investigation.

### Success Rates and Long-Term Outcomes

Our study confirms the findings of Kulkarni et al. [7], who reported that ETV success is highly dependent on patient selection, particularly favoring children older than 1 year with aqueduct stenosis. Similarly, our meta-regression analysis found that larger sample sizes were associated with improved outcomes for ETV, reinforcing that studies with limited patient numbers may overestimate VPS success rates due to selection bias.

The Cochrane Review [13], a widely cited meta-analysis, concluded that there was insufficient high-quality evidence to support ETV over VPS as the preferred treatment, largely due to variability in patient selection and follow-up duration. Our findings partially align with this conclusion, demonstrating that VPS has a higher short-term success rate but carries a greater long-term failure risk due to shunt dependency and infections. Unlike previous meta-analyses, however, we incorporated heterogeneity assessment (I^2^ = 22.5%) and meta-regression modeling, which allowed for a more detailed examination of study-level factors affecting outcomes.

### Age-Dependent Variability in Outcomes

Several studies, including Greuter et al. [23], have demonstrated that ETV has a lower success rate in infants under 6 months of age, due to immature cerebrospinal fluid (CSF) absorption pathways and a higher risk of ETV failure within the first year. In our review, studies that included a higher proportion of younger infants [16–18] reported higher VPS success rates, which may reflect the reduced efficacy of ETV in this age group. However, studies focused on older children [17,18] showed ETV performed significantly better in reducing shunt dependency.

This variability highlights the importance of stratified analyses when comparing treatment options. The ETV Success Score (ETVSS), as proposed by Kulkarni et al. (2009), is a critical tool that should be applied more consistently across studies to improve the comparability of findings.

### Complication Rates: Infection and Shunt Dependency

Consistent with prior research, our findings reinforce that VPS has a significantly higher infection rate compared to ETV. Tuli et al. [24] found that VPS infections occur in 10–20% of cases within the first year, with a substantial proportion requiring shunt revisions. Our meta-analysis aligns with this, as studies that included longer follow-up periods [19–21], reported higher VPS failure rates due to infections and mechanical malfunctions.

However, ETV is not without risks. Studies identified intraoperative bleeding, CSF leakage, and late failure as key complications. In our analysis, studies with higher risk of bias [15–20] tended to underreport complications, suggesting possible measurement bias in ETV studies.

### Long-Term Neurocognitive Outcomes

Several studies have examined the neurocognitive outcomes of patients undergoing ETV versus VPS. Their findings suggest that ETV patients demonstrate better long-term cognitive function, likely due to the avoidance of shunt-related complications and the preservation of normal CSF physiology. Our meta-analysis supports this conclusion, as studies with longer follow-ups [16,18] reported better developmental outcomes in ETV-treated children. However, these benefits were more pronounced in older children, reinforcing the age-dependent variability of treatment success.

### Heterogeneity and Study-Level Biases

One of the major contributions of this review is the incorporation of heterogeneity assessment and bias analysis. Previous systematic reviews often failed to account for between-study variability, leading to inconsistent conclusions. Our findings suggest that:

- Studies with high sample sizes and longer follow-up periods [13,16] showed greater reliability.
- Studies with small sample sizes and high risk of bias [14] tended to favor VPS, possibly due to selection bias or shorter follow-up durations.
- Meta-regression analysis showed that risk of bias negatively correlated with ETV success (β = −0.065, p = 0.12), suggesting that studies with poor methodology may underestimate ETV benefits.

## Conclusion

This study expands upon previous research by incorporating heterogeneity analysis, meta-regression modeling, and risk of bias assessments to provide a more comprehensive comparison of ETV and VPS in pediatric hydrocephalus. While our findings align with past studies demonstrating ETV’s superiority in reducing long-term shunt dependency and infections, they also highlight age-dependent variability and the need for standardization in patient selection criteria. Future research should focus on randomized controlled trials with longer follow-up periods to eliminate confounding variables and provide stronger clinical guidance.

This study highlights the advantages and drawbacks of both ETV and VPS, reinforcing ETV as a viable alternative to shunting in appropriately selected patients. However, methodological inconsistencies and study-level biases necessitate higher-quality, randomized controlled trials to strengthen clinical recommendations.

## Data Availability

All files from the systematic review and meta-analysis are available in our own databases, which are not published.

